# Temperature Significantly Change COVID-19 Transmission in 429 cities

**DOI:** 10.1101/2020.02.22.20025791

**Authors:** Mao Wang, Aili Jiang, Lijuan Gong, Lina Lu, Wenbin Guo, Chuyi Li, Jing Zheng, Chaoyong Li, Bixing Yang, Jietong Zeng, Youping Chen, Ke Zheng, Hongyan Li

**Author notes:** Corresponding author: Name: Mao Wang Ph.D., Associate Prof, Deparment of Occupatinal and Environmetal Health, School of Public Health, Sun Yat-sen University (Northern Campus), 74 Zhongshan Road II Guangzhou 510080 PR-China, (M Wang).

## Abstract

**Background:** There is no evidence supporting that temperature changes COVID-19 transmission.

**Methods:** We collected the cumulative number of confirmed cases of all cities and regions affected by COVID-19 in the world from January 20 to February 4, 2020, and calculated the daily means of the average, minimum and maximum temperatures in January. Then, restricted cubic spline function and generalized linear mixture model were used to analyze the relationships.

**Results:** There were in total 24,139 confirmed cases in China and 26 overseas countries. In total, 16,480 cases (68.01%) were from Hubei Province. The lgN rose as the average temperature went up to a peak of 8.72°C and then slowly declined. The apexes of the minimum temperature and the maximum temperature were 6.70°C and 12.42°C respectively. The curves shared similar shapes. Under the circumstance of lower temperature, every 1°C increase in average, minimum and maximum temperatures led to an increase of the cumulative number of cases by 0.83, 0.82 and 0.83 respectively. In the single-factor model of the higher-temperature group, every 1°C increase in the minimum temperature led to a decrease of the cumulative number of cases by 0.86.

**Conclusion:** The study found that, to certain extent, temperature could significant change COVID-19 transmission, and there might be a best temperature for the viral transmission, which may partly explain why it first broke out in Wuhan. It is suggested that countries and regions with a lower temperature in the world adopt the strictest control measures to prevent future reversal.

## Introduction

For the COVID-19 outbreak, it is important to understand its biological characteristics in the natural environment, especially during the transmission. Temperature could be an important factor that exerts different impact on people’s living environment in different parts of the world under different climate conditions, which can play a significant role in public health in terms of the epidemic development and control.^1^

The first COVID-19 case was admitted to the hospital on December 12, 2019, and the first generation of patients was identified on January 6, 2020. It was on January 20 when secondary generation were confirmed that human-to-human transmission was considered possible.^2^ More recently since February 4, cases of the third- and fourth-generation transmission have been reported.^3^ However, we still don’t understand the best temperature for its transmission and what the range could be. The question remains how many cases would be added to the cumulative number of daily confirmed cases when temperature increases by 1°C.

The lack of knowledge can dilute the strength and focus of the prevention and control measures. Research on SARS revealed that the temperature in the four major affected cities including Beijing and Guangzhou was significantly related to the outbreak. Studies found that during the outbreak of SARS in 2003, when the temperature was low, the risk of increasing daily incidence rate could be 18.18 times higher than that under higher temperature.^4^ This finding could be a clue for us to understand the temperature-transmission relation of COVID-19 as it shares genetic similarities with SARS. However, there is no such study published so far.

Our hypothesis is that different temperature could significantly influence the transmission of the virus and nonlinear dose-response relationship exists between the two. We also speculate that there is certain temperature that best fit the benefit of the virus and that lower temperature contributes to the transmission. This could be closely related to Wuhan and its neighboring areas, the epicenter of the outbreak. We collected full-sample data from the cities and regions affected by the virus around the world and analyzed their respective average, minimum and maximum temperature values to see if significant relations exist and if relatively accurate dose-response relationship could be concluded.

## Methods

### Study population

The study populations are the daily confirmed newly cases of COVID-19 officially reported in China and overseas countries and calculate the number of cumulative total confirmed cases in all cities and regions from January 20 to February 4, 2020. The population data were collected from the reports released on the official websites of the Health Commissions at all levels in China and the health authorities of overseas countries. Thus, no ethical review was required.

### Average, minimum and maximum temperatures

The daily average, minimum and maximum temperatures of all sampled cities from January 1 to 30, 2020 were collected from the meteorological authority in China and in other countries (the data of the capital cities were used) to calculate the daily means of the average, minimum and maximum temperatures in January.

### Statistical analysis

Descriptive analysis was performed. Numerical variables were described with means and standard deviations, and categorical variables such as frequencies with medians or percentages. The mean value of the case numbers in the three days following the first diagnosed case (the day of the first case included) in these regions and cities were calculated, and the number was taken as a variable marked as Day3. Log-transformation was performed for the values of Day3 and the number of cumulative total confirmed cases in all cities and regions with log10^number^+1. R (restricted cubic spline function, the related function packages used were “ggplot”, “spline”, “rms” in R 3.5.1.) was used to calculate the relationships between the three types of temperature data and the number of cumulative total confirmed cases (lgN), respectively to obtain the fitting equation and splines. Then, the generalized linear mixed model in SAS (version 9.4; SAS Institute Inc.) was developed to analyze the dose-response relationship. Two models were established, Model 1 with single factor correlation and Model 2 with the variable lgDay3. P<0.05 was considered statistical significance.

## Results

There were in total 24,139 confirmed cases in 34 provinces (including municipalities, autonomous regions and special administrative regions) in China and 26 overseas countries. In total, 16,480 cases (68.01%) were from Hubei Province (Table 1). The data of confirmed cases of Guangdong, Zhejiang, Henan, Jiangxi and Hunan were 931, 947, 801, 550 and 619; The data of average temperature were 5.23(0.95),17.30(1.30),9.36(2.12),2.64(1.45),2.90(6.34),6.52(0.99) while the data of minimum temperature were 1.25 (2.74), 12.33 (2.97), 5.10 (2.80), −1.12 (1.82), −1.87 (7.34) and 3.05 (1.25); and the data of maximum temperature were 9.15 (2.69), 22.28 (2.24), 12.72 (3.45), 6.39 (1.77), 7.67 (6.15) and 9.15 (2.69). And the values of Day3 were 18.43 (19.08), 1.20 (0.86), 2.08 (1.54), 2.18 (1.98), 1.12 (0.91) and 1.64 (0.99) (Table 1). The cumulative number of confirmed cases, average temperature, minimum temperature, maximum temperature and Day3 of Wuhan were 8153, 4.47, 1.37, 7.57 and 75.70 respectively. The data of other cities in Hubei Province were listed in Table 1, and national map the five variables of all cities in China are showed in Supplemental Figure S1-S5.

**Table 1.** General characteristics of temperature and related variables with all cities and regions affected by COVID-19 in the world.

| Country/<br>Province | N/number<br>of cities | Average<br>Mean (SD) | Minimum<br>Mean (SD) | Maximum<br>Mean (SD) | Day3<sup>#</sup><br>Mean (SD) |
| --- | --- | --- | --- | --- | --- |
| China |  |  |  |  |  |
| Anhui | 462/16 | 4.58(1.27) | 1.40(1.43) | 7.75(1.79) | 1.10(0.82) |
| Beijing | 211/14 | -1.21(1.37) | -9.06(5.78) | 7.63(1.43) | 0.69(0.46) |
| Chongqing | 376/37 | 8.39(2.35) | 2.81(2.52) | 13.47(1.48) | 0.94(0.49) |
| Fujian | 207/9 | 15.11(1.25) | 11.07(1.56) | 19.15(1.39) | 1.74(1.27) |
| Gansu | 57/12 | -2.82(3.52) | -10.31(4.83) | 4.50(4.17) | 0.58(0.38) |
| Guangdong | 931/20 | 17.30(1.30) | 12.33(2.97) | 22.28(2.24) | 1.20(0.86) |
| Guangxi | 151/12 | 15.52(2.36) | 11.05(3.63) | 19.98(3.29) | 1.25(0.96) |
| Guizhou | 64/11 | 7.89(3.38) | 2.79(1.98) | 12.27(5.78) | 0.60(0.36) |
| Hainan | 109/15 | 21.07(2.47) | 13.69(3.02) | 28.59(2.19) | 1.16(1.01) |
| Hebei | 135/11 | -1.73(2.34) | -6.56(3.24) | 3.11(1.61) | 1.21(0.78) |
| Heilongjiang | 189/12 | -13.62(7.05) | -20.13(8.94) | -6.94(5.65) | 1.00 (0.82) |
| Henan | 801/20 | 2.64(1.45) | -1.12(1.82) | 6.39(1.77) | 2.18(1.98) |
| Hongkong | 16/1 | 18.00 | 11.00 | 25.00 | 1.33 |
| Hubei | 16480/17 | 5.23(0.95) | 1.25(2.74) | 9.15(2.69) | 18.43(19.08) |
| Hunan | 619/13 | 6.52(0.99) | 3.05(1.25) | 9.99(2.82) | 1.64(0.99) |
| Jiangsu | 341/13 | 5.23(1.61) | 1.17(2.48) | 9.29(3.45) | 1.08(0.78) |
| Jiangxi | 550/11 | 2.90(6.34) | -1.87(7.34) | 7.67(6.15) | 1.12(0.91) |
| Jilin | 54/8 | -11.82(1.50) | -18.58(2.53) | -5.06(1.79) | 0.46(0.25) |
| Liaoning | 157/13 | -4.44(4.36) | -11.15(4.21) | 0.27(2.67) | 1.28(0.76) |
| Macao | 15/1 | 17.50 | 10.00 | 25.00 | 0.67 |
| Neimenggu | 47/12 | -2.80(3.75) | -10.13(4.45) | -17.47(6.56) | 0.64(0.22) |
| Ningxia | 34/6 | -0.17(4.82) | -11.83(1.83) | 6.67(0.82) | 0.55(0.55) |
| Qinghai | 17/2 | -8.66(3.45) | -15.07(4.95) | -2.25(1.95) | 1.34(0.94) |
| Shaanxi | 143/12 | 1.24(2.70) | -3.28(3.83) | 5.77(2.35) | 0.83(0.66) |
| Shandong | 298/15 | 1.95(1.44) | -1.68(2.27) | 5.59(1.65) | 1.67(0.62) |
| Shanghai | 110/15 | 10.08(1.56) | 2.35(2.83) | 18.11(4.96) | 1.37(1.35) |
| Shanxi | 80/11 | 3.23(2.39) | -2.28(3.68) | -7.78(5.10) | 0.85(0.62) |
| Sichuan | 164/15 | 5.74(6.16) | 1.82(9.74) | 9.67(3.02) | 1.26(1.42) |
| Taiwan (Taibei city) | 16/1 | 16.00 | 9.00 | 23.00 | 0.33 |
| Tianjin | 66/13 | 0.92(2.01) | -6.77(2.52) | 7.85(2.30) | 0.92(0.87) |
| Xinjiang | 20/8 | -11.97(4.46) | -18.46(5.56) | -5.49(4.19) | 0.54(0.35) |
| Xizang (Lhasa city) | 1/1 | -3.50 | -20.00 | 13.00 | 0.33 |
| Yunnan | 122/14 | 11.97(3.30) | 5.73(3.35) | 18.35(3.66) | 0.85(0.42) |
| Zhejiang | 947/12 | 9.36(2.12) | 5.10(2.80) | 12.72(3.45) | 2.08(1.54) |
| Other countries |  |  |  |  |  |
| Australia | 14/1 | 13.00 | 20.40 | 27.70 | 1.33 |
| Belgium | 1/1 | -0.10 | 2.50 | 5.10 | 0.33 |
| Brazil | 1/1 | 17.40 | 22.20 | 26.90 | 0.33 |
| Cambodia | 1/1 | 21.70 | 26.40 | 31.10 | 0.33 |
| Canada | 4/1 | -15.10 | -10.80 | -6.40 | 0.67 |
| Finland | 1/1 | -10.30 | -7.20 | -4.10 | 0.33 |
| France | 6/1 | 1.10 | 3.60 | 6.10 | 1.00 |
| German | 12/1 | -2.90 | -0.60 | 1.80 | 1.66 |
| Indian | 3/1 | 7.60 | 14.30 | 21.00 | 0.33 |
| Italy | 2/1 | 1.90 | 7.00 | 12.10 | 0.67 |
| Japan | 20/1 | 1.20 | 5.40 | 9.50 | 1.00 |
| Korea | 15/1 | -6.10 | -2.80 | 0.60 | 0.67 |
| Malaysia | 10/1 | 22.80 | 27.30 | 31.70 | 1.33 |
| Nepal | 1/1 | 2.00 | 10.00 | 18.00 | 0.33 |
| Philippines | 3/1 | 21.70 | 25.90 | 30.00 | 1.00 |
| Russian | 2/1 | -18.00 | -14.00 | -10.00 | 0.67 |
| Singapore | 27/1 | 23.10 | 26.50 | 29.90 | 1.33 |
| Spain | 1/1 | 0 | 5.30 | 10.60 | 0.33 |
| Sri Lanka | 1/1 | 23.30 | 26.70 | 30.00 | 0.33 |
| Sweden | 1/1 | -5.00 | -2.90 | -0.70 | 0.33 |
| Thailand | 23/1 | 21.70 | 26.70 | 31.70 | 0.67 |
| United Arab Emirates | 5/1 | 14.40 | 18.60 | 22.80 | 0.33 |
| United Kingdom | 2/1 | 2.20 | 5.00 | 7.80 | 0.67 |
| USA | 10/1 | -1.70 | 2.20 | 6.10 | 0.67 |
| Vietnam | 10/1 | 14.40 | 16.70 | 18.90 | 0.67 |

<sup>#</sup> The mean value within three days from the first confirmed cases

Simulation equation analysis was conducted with lgN and the average, minimum and maximum temperature values respectively, and all results indicate significant correlation between the two (p<0.0001). In the equation of the average temperature and lgN, as shown in Figure 1a when X_0_=8.72°C, lgN was 2.0949 and the corresponding cumulative number of cases was 12.44. In the equation of the minimum temperature when X_0_=6.70, lgN was 2.0991 and the corresponding cumulative number of cases was 12.56 (Figure 2a). In the equation of the maximum temperature when X_0_=12.42, lgN was 2.0925 and the cumulative number of cases was 12.37 (Figure 3a). The inflection point temperature value was consistent with the range of temperature in Hubei Province including Wuhan, which the range of average, minimum and maximum temperature are 4.28∼6.18, −2.51∼3.99, 6.46∼11.84 (Table 1). The analysis showed that lgN increased as the average temperature rose and started to decline slowly when X_0_ reached the apex (Figure 1b). The curves of the minimum and maximum temperatures were similar (Figure 2b and 3b). The difference lay in the fact that the minimum temperature curve increased rapidly but declined at the slowest rate, while the maximum temperature curve had almost the same increase and decrease rates.

**Figure 1a.**
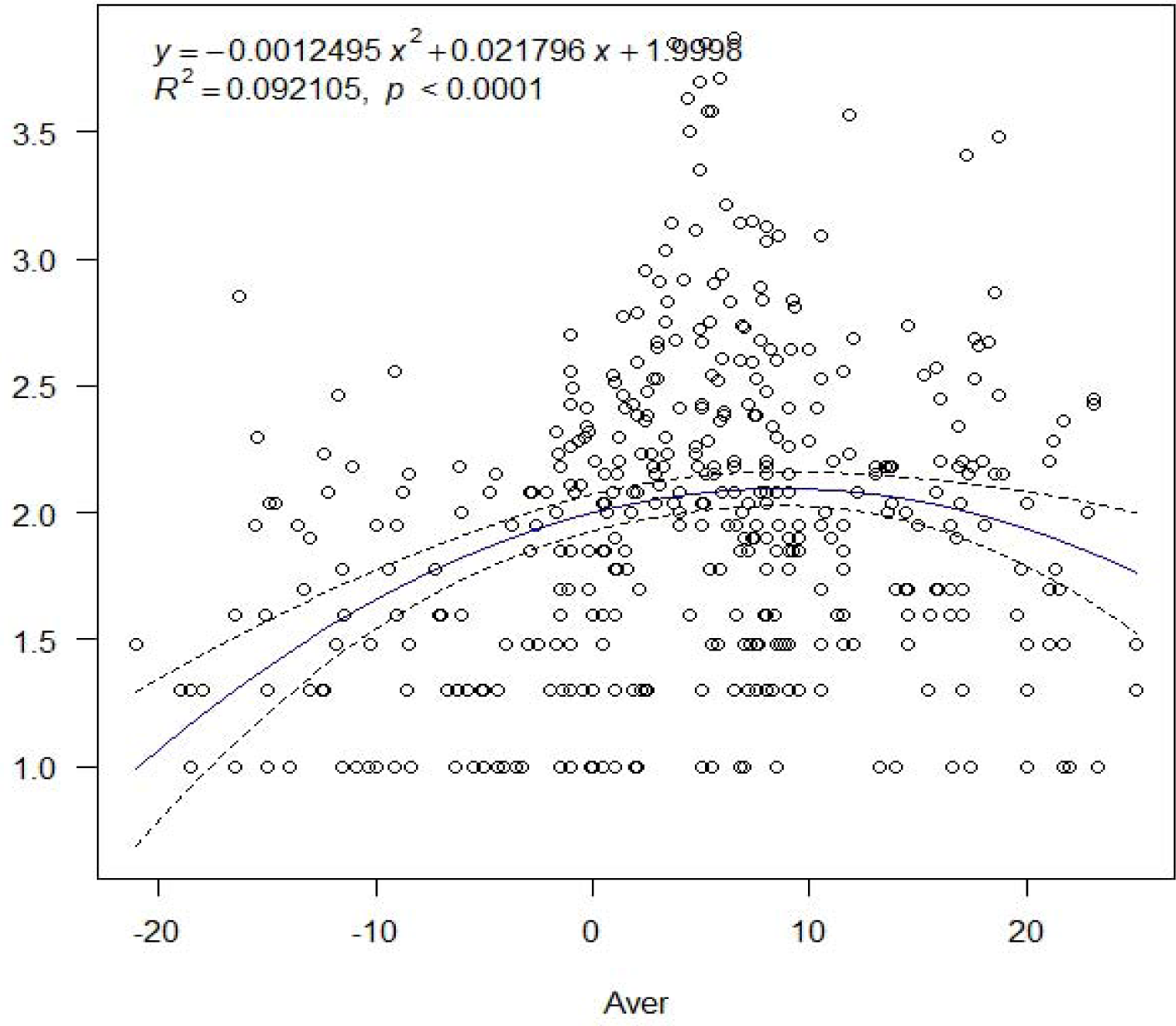
The relationship between average temperature and lgN of COVID-19 transmission in the world.

**Figure 1b.**
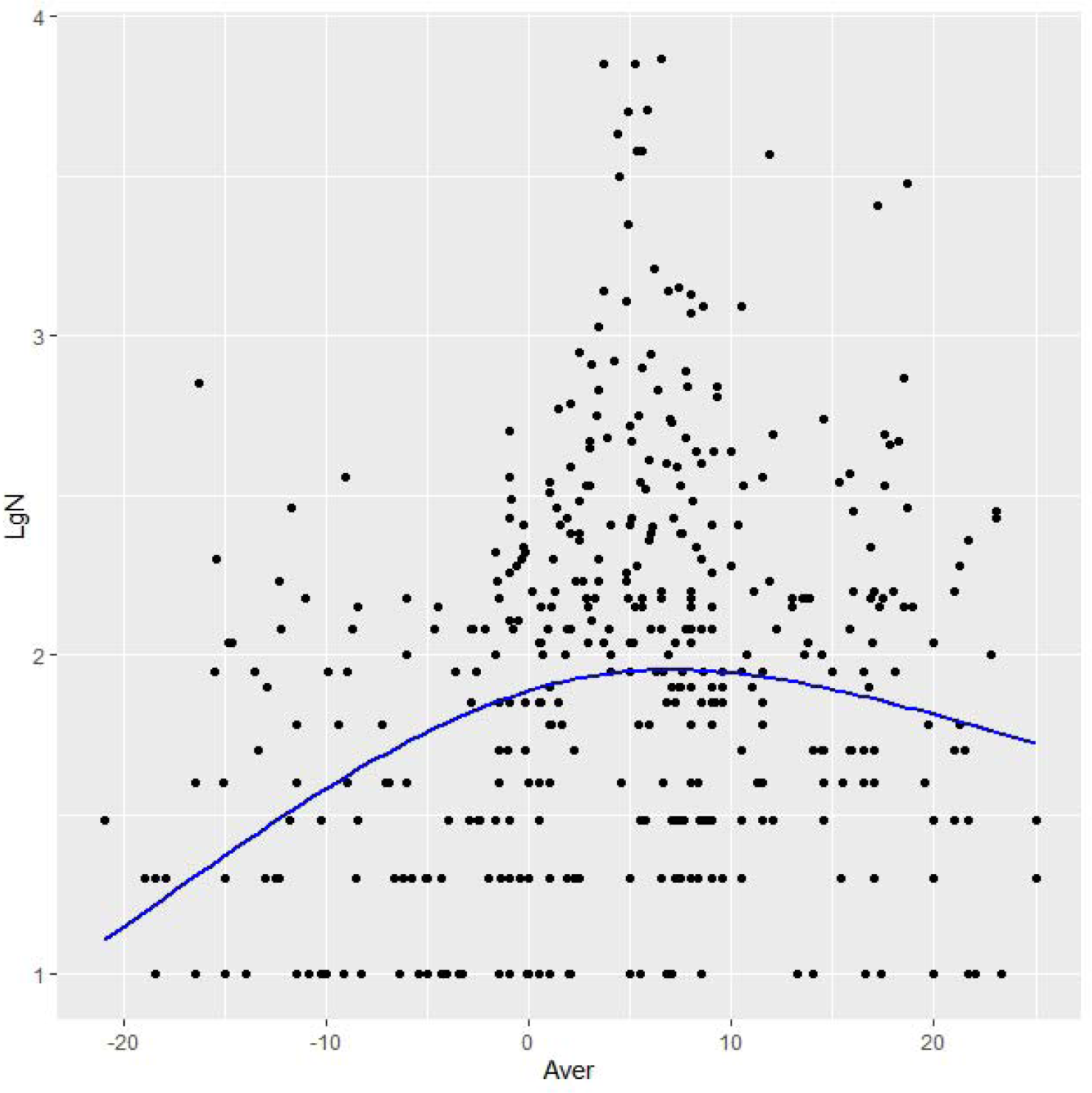
The cubic spline curve between average temperature and lgN of COVID-19 transmission in the world.

**Figure 2a.**
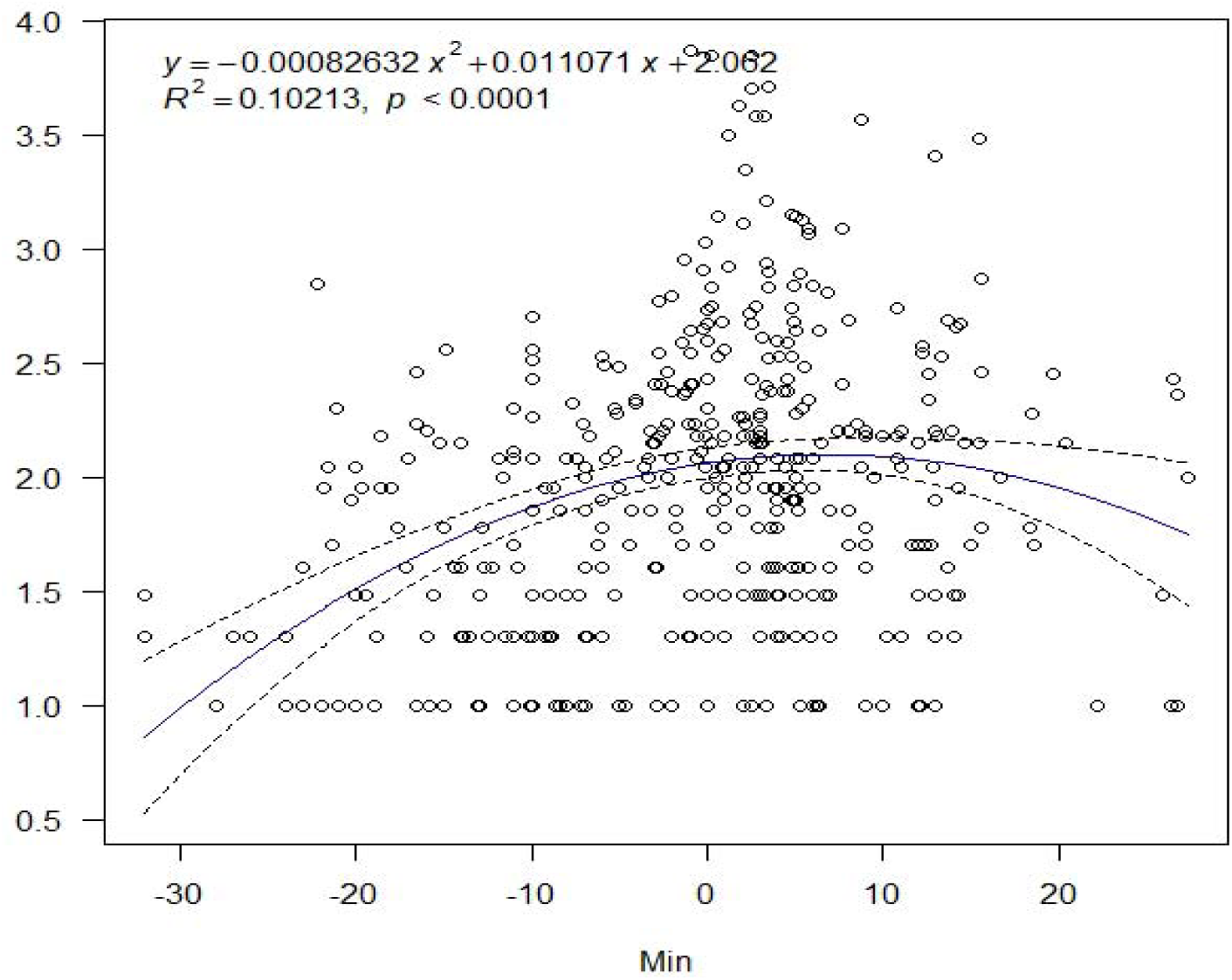
The relationship between minimum temperature and lgN of COVID-19 transmission in the world.

**Figure 2b.**
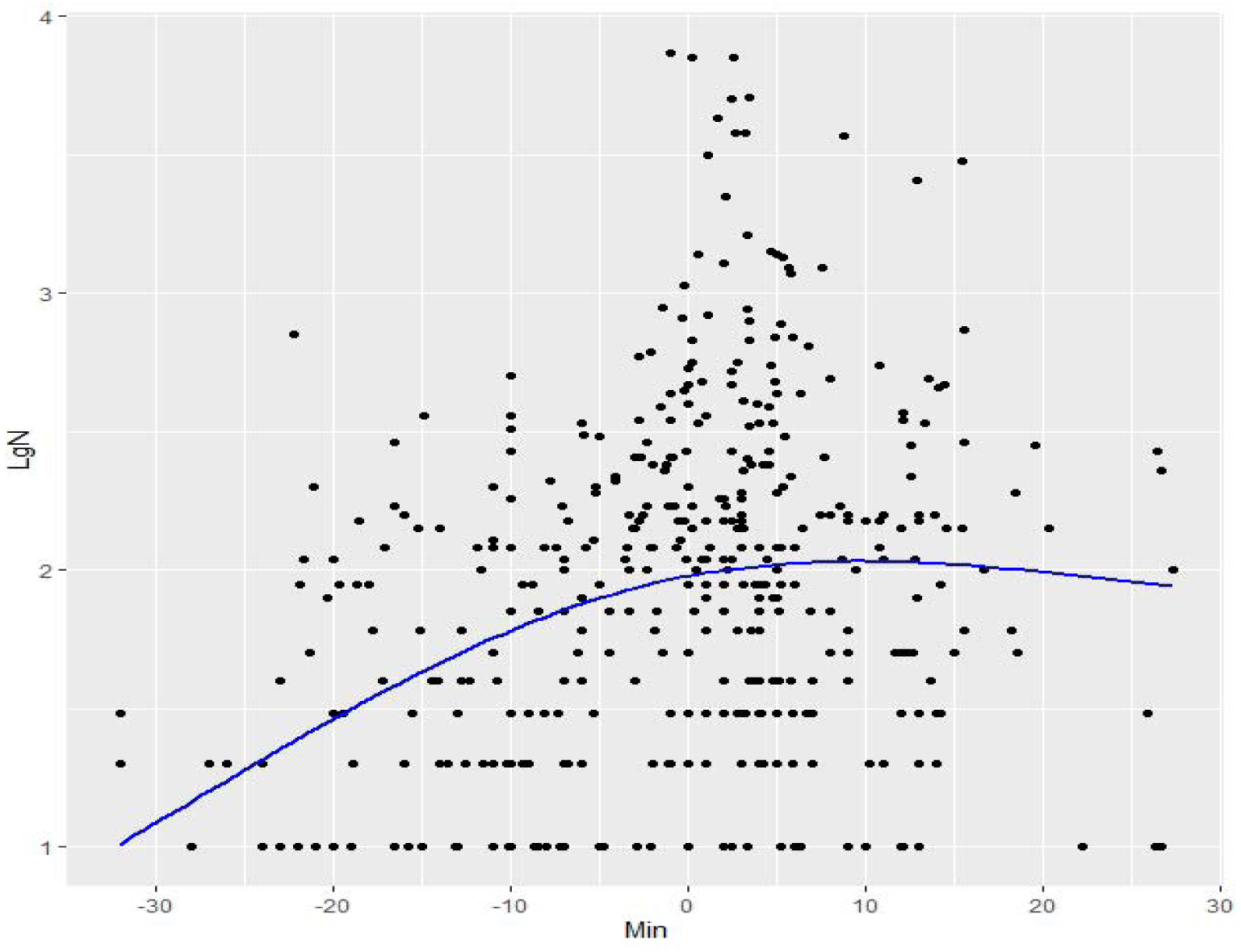
The cubic spline curve between minimum temperature and lgN of COVID-19 transmission in the world.

**Figure 3a.**
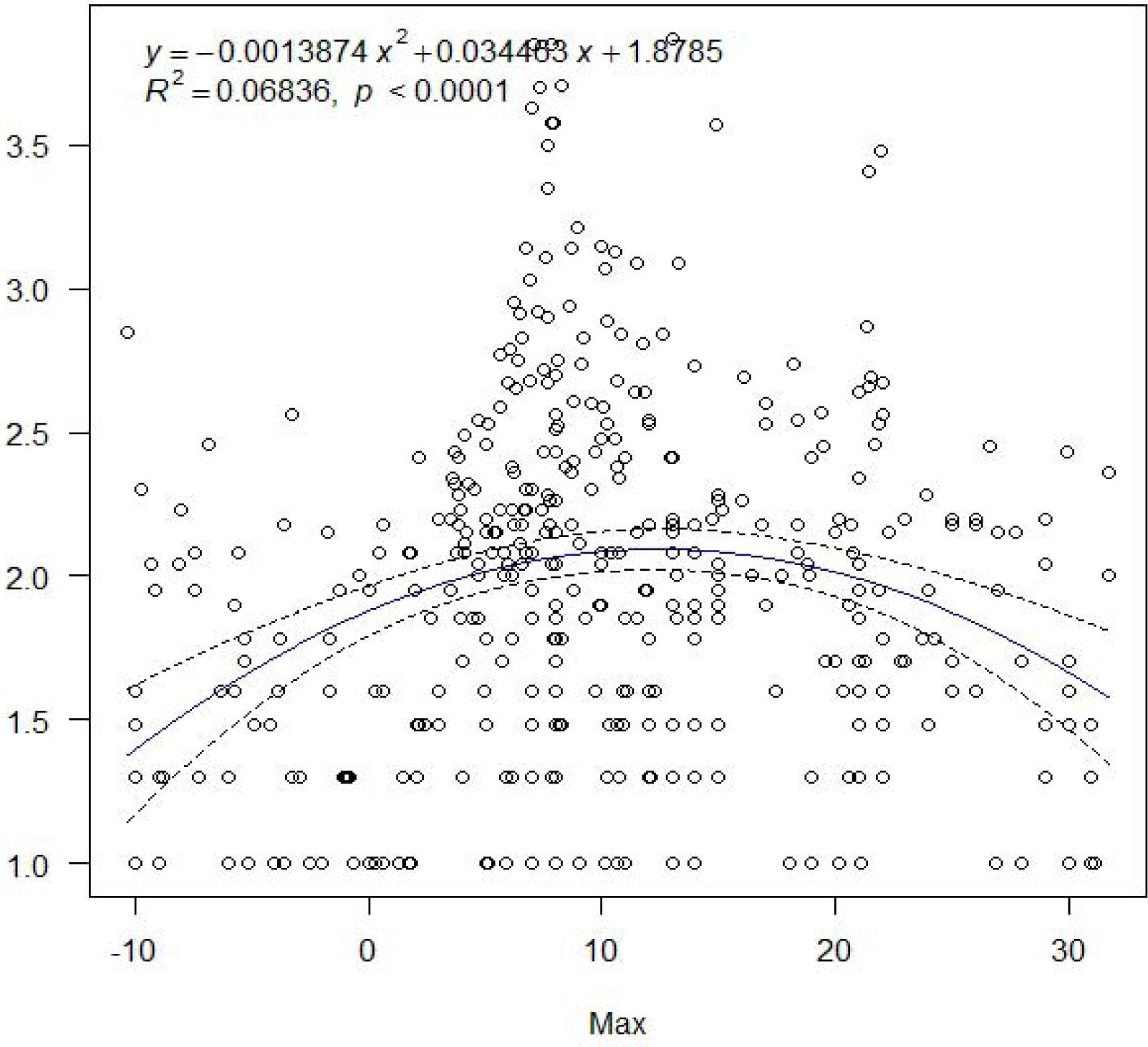
The relationship between maximum temperature and lgN of COVID-19 transmission in the world.

**Figure 3b.**
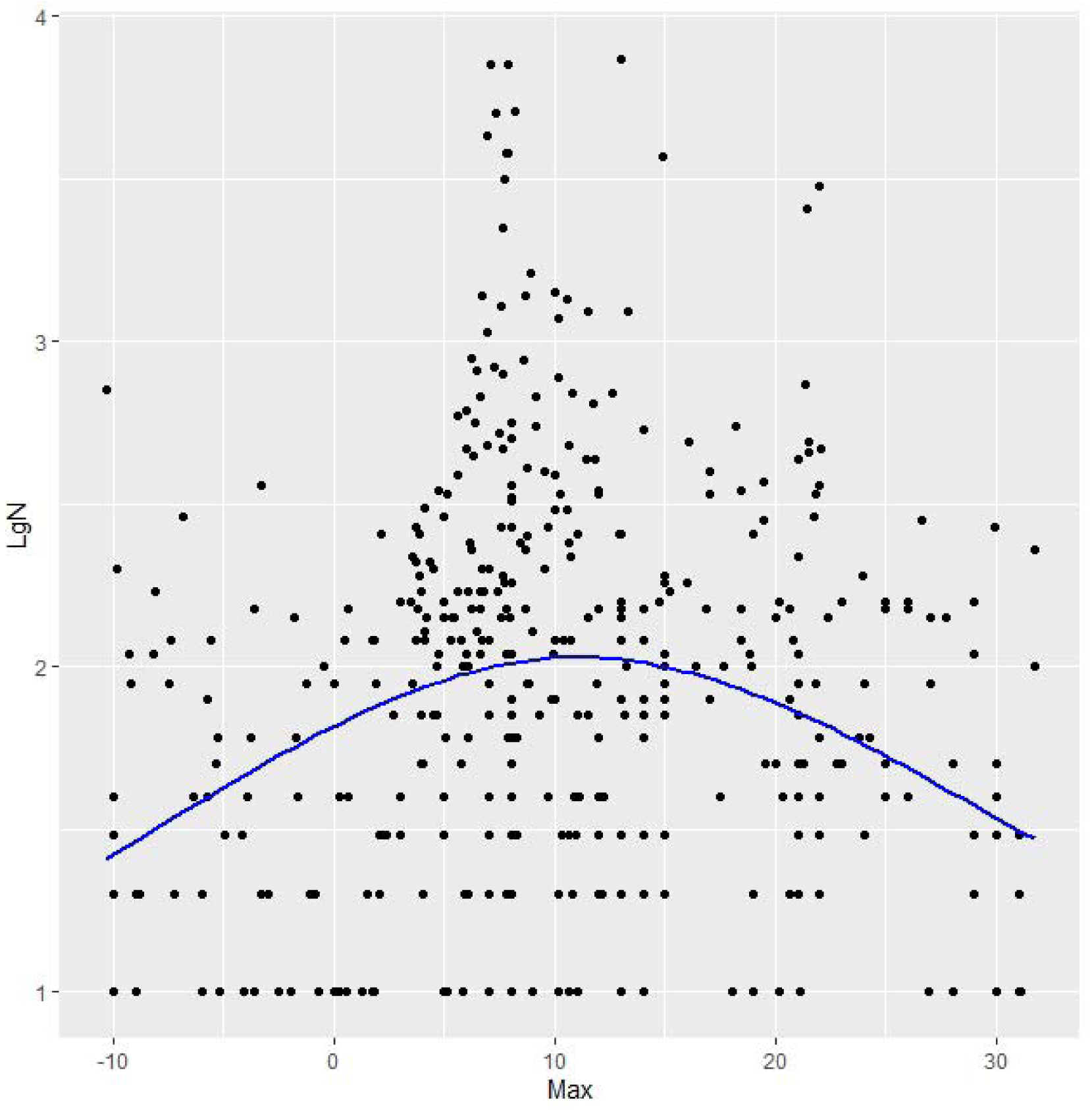
The cubic spline curve between maximun temperature and lgN of COVID-19 transmission in the world.

Segmentation points were defined according to X_0_ in the equation and divided into cities with lower temperature and cities with higher temperature. And the generalized linear regression model was then used for further analysis. Model 1 was a single factor regression, and model 2 included the variable lgDay3. In the models of the lower-temperature group, the three types of temperature data demonstrated significant correlation with lgN (Table 2). In the higher-temperature group, only the minimum temperature data showed significant correlation with lgN in model 1, while such correlation disappeared in model 2. In the lower-temperature group, every 1°C increase in the average, minimum and maximum temperatures in model 1, lgN increased by 0.036, 0.028 and 0.032 respectively; and the corresponding cumulative number of case by 0.86, 0.85 and 0.86. In model 2, lgN increased by 0.018, 0.014 and 0.020, and the corresponding cumulative number of cases by 0.83, 0.82 and 0.83 (Table 2).

**Table 2.** The dose-response relationship between temperature and the cumulative number of cases in 429 cities and regions by a generalized linear mixed model.

|  | LgN<sup>□</sup> |  |  |  |
| --- | --- | --- | --- | --- |
|  | Model 1 |  | Model 2<sup>#</sup> |  |
|  | Estimated (95% CI) | P value | Estimated (95% CI) | P value |
| Average |  |  |  |  |
| Higher | -0.015(-0.036,-0.007) | 0.178 | -0.011(-0.028,0.007) | 0.231 |
| LgDay3<sup>*</sup> |  |  | 0.991(0.746,1.235) | <0.0001 |
| Average |  |  |  |  |
| Lower | 0.036(0.026,0.045) | <0.0001 | 0.018(0.011,0.025) | <0.0001 |
| LgDay3<sup>*</sup> |  |  | 1.159(1.037,1.282) | <0.0001 |
| Minimum |  |  |  |  |
| Higher | -0.007(-0.033,0.018) | 0.563 | 0.006(-0.025,0.013) | 0.550 |
| LgDay3<sup>*</sup> |  |  | 1.142(0.868,1.416) | <0.0001 |
| Minimum |  |  |  |  |
| Lower | 0.028(0.021,0.036) | <0.0001 | 0.014(0.008,0.019) | <0.0001 |
| LgDay3<sup>*</sup> |  |  | 1.115(0.993,1.236) | <0.0001 |
| Maximum |  |  |  |  |
| Higher | -0.021(-0.038,-0.005) | 0.011 | -0.006(-0.018,0.007) | 0.361 |
| LgDay3<sup>*</sup> |  |  | 1.120(0.915,1.325) | <0.0001 |
| Maximum |  |  |  |  |
| Lower | 0.032(0.019,0.046) | <0.0001 | 0.020(0.011,0.029) | <0.0001 |
| LgDay3<sup>*</sup> |  |  | 1.173(1.043,1.302) | <0.0001 |

<sup>#</sup> Model 2: model 1+lgDay3 <sup>\*</sup>The mean value within three days from the first confirmed cases
□ $\log_{10}^{\text{number}} + 1$

In the higher temperature group, every 1°C increase in the minimum temperature in model 1, lgN decreased by 0.068 and the corresponding cumulative number of cases decreased by 0.86; and lgDay3 demonstrated significant correlations in all model 2 analysis in this group (p<0.0001) (Table 2).

## Discussion

Significant impact of different temperature exposure on the human-to-human transmission of COVID-19 was first identified through our study. It is also found that there exists nonlinear dose-response relationship, which means that a best temperature might exist for the transmission and that lower temperature contributes to the growth and transmission of the virus. The fact that the outbreak emerged in Wuhan and its neighboring areas could be closely related to the temperature of the local region. The results indicate that strict public health strategies shall be continued when temperature would drop in most part of the country so as to prevent reversal of the epidemic.

Human beings noticed the close relationship between epidemics and seasons as early as in ancient Greece.^5^ For example, in temperate regions, May to September in the southern hemisphere and November to March in the northern hemisphere is the high season for influenza. Academic journals including *Nature* started the discussed on the airborne transmission of influenza virus in low-temperature, low-humidity environment in Mid-20^th^ Century.^6^ After years of research, low temperature, low-humidity environment is now generally recognized as the key factor contributing to the transmission of influenza in winter.^1,7^ In addition to epidemiological statistics, many researches had also shown more direct lab evidence.^5,8-10^ In a study on swine flu performed in 2007 in a strictly controlled indoor environment other factors, researchers found that, after they excluded the possibility of immune system impact from the low temperature, the virus could be transmitted through aerosol.^11^ The study also found that the infectivity of the virus was stronger under lower-temperature, lower-humidity conditions. The infection rate was 75-100% in an environment where temperature was 5°C and relative humidity 35% and 50%. When the temperature was increased to 30°C and the relative humidity was 35%, the infectious rate was 0. These above results support our research. Based on our study, it is estimated that when the temperature reaches 30°C, the cumulative number of cases would only increase by 3.38. This indicates that the virus is highly sensitive to high temperature that would prevent the virus from spreading.

Currently, there is no study on the impact of temperature and humidity on the transmission of the COVID-19. But a study reviewed the outbreak of SARS in four affected cities in China including Guangzhou and Beijing and pointed out the significant correlation between the temperature and virus transmission.^4^ The paper identified three key factors contributing to SARS transmission, namely, temperature, humidity and wind speed.^12^ An article published in 2011 conducted a lab experiment on SARS and found that the virus could maintain active for at least five days on smooth surface in an environment where temperature was 22-25°C and relative humidity 40-50%.^13^ When the temperature increased to 38°C and the relative humidity to 95%, the virus soon lost its activity. Another virus sharing genetic similarities with COVID-19 is MERS; a study published in 2013 found that MERS could maintain its activity for a long time both as droplets on solid surface and as aerosol as long as in low-temperature, low-humidity environment.^14^ Researchers also performed experiments on other coronaviruses (excluding SARS and MERS), and the same conclusions were drawn.^15,16^ Thus, in general, low temperature and low humidity significantly contribute to the transmission and survival of coronaviruses. All the findings cited support the direct link between virus transmission and the temperature and these above results support with our findings.

In our study, the cumulative number of confirmed COVID-19 cases from 429 cities and regions (data of capital cities were adopted for the national-level fitting curves) across the world were collected to analyze the impact of the three types of temperature data. The results provide relative sound evidence to the dose-response relationship between temperature and the transmission of the virus. The dose-response relationship conclusion was obtained from the analysis of data collected under the circumstances where strong, effective control measures had been adopted by the government and CDCs at all levels in China as well as healthcare professionals. The valuable results show that temperature can exert relatively big impact on the transmission of the disease, indicating close relationship between the temperature and the emergence and spread of the epidemic in Wuhan and its neighboring areas. It also indicates that the temperature drop in recent days in China might pose important influence on possible reversal of the epidemic.

The time period from January 20 to February 4, 2020 was defined for the epidemiological data analysis for four reasons. First, the longest incubation period was reported to be 14 days. The time period covers the first 14 days after large-scale human-to-human transmission was first identified outside Wuhan.^2,3^ Second, epidemiological evidence indicated potential human-to-human transmission on January 20. Though lockdown of Wuhan and seven neighboring cities was implemented at 10:00 am on January 23, the transportation was not fully cut off, which allowed an outflow of nearly 5 million people from the region.^17^ Third, the Chinese government adopted the strictest control measures on February 4 to control the movement of people and discourage gathering in enclosed or semi-enclosed public areas such as restaurants. Fourth, a decrease in case numbers was indicated on the curve of Beijing for five consecutive days since February 4.^18^ Therefore, the time period from January 20 to February 4 was of analytic importance for this study.

The reasons that January 1 to 30, 2020 was selected to calculate the daily temperature mean values are threefold. First, based on an incubation period of 3-14 days and the time period for case data analysis in this study (January 20 to February 4), January 30, four days prior to February 4, was defined as the endpoint because at least 4 days was required for case confirmation with the shortest incubation period (3 days of incubation and 1 day for lab diagnosis and case reporting); and January 1 was determined as the start point to include the cases with the longest incubation period (14 or 24 days). Second, the Chinese government initiated the strictest measures on February 4, which minimized the movement of people and limited their activities at home. This changed significantly the environment and the transmission pattern of the disease as well as the probability of exposure and infection. Third, the traditional Chinese New Year and the winter vacation of schools and universities fell on January this year, which means frequent movement of travelers in large numbers with a potential of creating billions of travels.

This study adopted the numbers of daily newly confirmed cases officially released across the world and full-sample data in China for analysis, and defined the key epidemiological evaluation time period (January 20 to February 4) as well as the key time period of temperature exposure (January 1 to 30) to calculate the daily means of three types of temperature data in January. A total of one million entries of daily temperature data released by the meteorological authorities covering the countries and regions with a total population around 2 billion were collected for the time-space analysis. The new variable Day3 was defined for the analysis as a control to evaluate the impact of the number of imported confirmed cases on the dose-response relationship.

There are several limitations of the study. First, this is a time-space cross-sectional study. Thus, no causal relationship can be proved. But the nonlinear dose-response relationship was concluded. Second, official data regarding the imported cases could not be obtained, making it impossible to analyze the impact of the imported case numbers on the exponential function. However, a new variable, Day3, was adopted for the control analysis. Third, the impact of temperature on sex and age could not be analyzed because the key information was not available on the official release in many cities except for Hong Kong SAR. Thus, the cumulative number of cases calculated based on the daily confirmed cases was used. Epidemiologically, the impact of temperature varies between sexes and among different age groups and has obvious influence on the middle-aged and the aged populations. Despite of the limitations, the outcome of this study demonstrates statistical significance, consistency and novelty and has led to relatively clear conclusions.

## Conclusion

The study concludes that temperature has significant impact on the transmission of COVID-19. There might be nonlinear dose-response relationship between the two, indicating that there is a best temperature contributing to its transmission and that low temperature is beneficial to the viral transmission. The emergence of the outbreak in Wuhan and its neighboring areas may be closely related to the local temperature. For countries and regions with a lower temperature, strict prevention and control measures should be continued to prevent future reversal of the epidemic.

## Data Availability

All the data in the manuscript are from the official website. Since this study included data from 403 cities and regions in China, we only added data links for some of them.

http://wjw.beijing.gov.cn/

http://wsjkw.sh.gov.cn/index.html

http://www.gzcdc.org.cn/

http://www.szcdc.net/rdzt/xxgzbd/

https://m.tianqi.com/

